# Research Paradigm of the International Classification of Functioning, Disability and Health (ICF) with Item Response Theory: Clarification, Classification, and Challenge

**DOI:** 10.1101/2023.05.14.23289950

**Authors:** Chun Feng, Shou-Guo Liu, Feng Lin

## Abstract

**Objectives:** The purpose of this literature review was to clarify the sufficiency and efficiency needs, as well as the stratification and the assignment scoring principles of the International Classification of Functioning, Disability and Health (ICF) core set, in conjunction with the Item Response Theory (IRT)-derived functioning outcome measures.

**Methods:** A literature search has been done on the PubMed database. We included studies involving the IRT-derived ICF evaluation tool. Our main outcome was the assignment and stratification scoring principle of IRT-derived ICF-based functioning outcome measures.

**Results:** 46 relative articles met the eligibility criteria of our review. On this basis, this review was undertaken to track the research progress of the ICF-based and IRT-modeling measurement tools and screen the final core set. This review included a classification of IRT modeling for ICF studies and a summary of existing research paradigms of IRT-derived ICF outcome measures. Moreover, this review identified blind spots related to scoring assignment principles and “difficulty parameter” in current ICF-based IRT studies.

**Conclusion:** The ICF is the worldwide terminology system for functioning categories, while ICF-based evaluation in clinical practice and scientific research has elicited the underlying challenges. However, the algorithm of the ICF-based IRT modeling may advance the understanding of ICF clinical application and enable a new paradigm for the design of IRT-derived ICF questionnaires, namely the parsimonious core set of ICF. Additionally, the Wright map has the potential to facilitate an understanding of the rehabilitation process and tailor rehabilitation goals.

## Introduction

Rehabilitation science is a discipline dedicated to comprehending and facilitating human functioning. In order to guide and inform clinical decisions, it is crucial to identify potential disabilities and monitor changes in functioning across the continuum of care. However, inconsistent and numerous outcome measure scales pose a major challenge to multidisciplinary communication. Additionally, determining the most appropriate functioning assessment scale presents several challenges. Firstly, it is inevitable to encounter a trade-off between the sufficiency and efficiency of assessment tools. Relying solely on disease-specific scales may fail to provide a comprehensive functioning assessment for patients with multiple health issues. A thorough assessment can provide valuable insights into rehabilitation management and functional recovery, while efficient clinical assessment tools that can rapidly and conveniently achieve outcome measures(Rolstad, Adler, and Rydén, 2011). The ideal functioning assessment should consider the length and complexity of assessment tools limited to a certain extent, so as to minimize the response burden on frail patients and optimize resource efficiency. Another concern is that many rehabilitation assessment scales, such as manual muscle testing, Barthel index, etc. use ordinal scales, that may not precisely capture the full range of functioning changes over time. To tackle these barriers, the World Health Organization has introduced the International Classification of Functioning, Disability and Health (ICF), which offers a standardized international terminology for defining and operationalizing function management across multiple domains. Accurate and timely assessment of an individual’s function is crucial in making clinical decisions for treatment planning and rehabilitation(Finch, Higgins, Wood-Dauphinee, and Mayo, 2009). To achieve this, the assessment scale must include items that have high discrimination, enabling the determination of functioning performance. The Item Response Theory (IRT) is a well-established psychometric paradigm that measures the interaction between examinees and test items to estimate an individual’s ability level (represented by the latent trait, θ). Additionally, the IRT model is a statistical technique that can provide theoretical and algorithmic support for the developing clinical assessment tools in healthcare practice(Stucki, Rubinelli, and Bickenbach, 2020), which can transform raw ICF scores into the IRT-based interval scores. Recently, emerging studies have advocated the use of IRT-based Rasch models to process ICF evaluation data and generate the new version of the ICF core set that contains the most discriminable and minimal items.

This review aims to offer an overview of the research paradigm regarding the ICF-based functioning evaluation in combination with the IRT technique tools. Incorporating these two techniques, a comprehensive and continuous assessment of human functioning with a minimal core set can be constructed for patients undergoing rehabilitation. It also explores the challenges of operationalizing and analyzing the ICF core set assessment, specifically in terms of stratification and assignment scoring principles. This review emphasizes the value of IRT modeling in ICF studies and further categorizes IRT-derived ICF outcome measures according to the scoring principles. Moreover, the review proposes appropriate assignment principles that match the difficult parameter in IRT modeling and discusses the benefits of using Wright maps to visualize rehabilitation goals and potential functioning barriers. Overall, this review focuses on introducing a technique-solid and framework-sound questionnaire construction approach to develop the parsimonious core set of ICF and the Wright map that can enhance clinical decision-making in rehabilitation settings.

## Materials and Methods

## Review Criteria

This review focused on studies that explore the psychometric properties of the IRT-derived ICF evaluation tool. The search was carried out in January 31^st^ 2023 based on the PubMed database. The keywords used during the search included the following: [(Rasch) AND (International Classification of Functioning, Disability and Health OR ICF), [(Item response theory OR IRT) AND (International Classification of Functioning, Disability and Health OR ICF)], and [(Mokken) AND (International Classification of Functioning, Disability and Health OR ICF)]. The reference lists of the resulting articles were scanned to identify further relevant studies, which totally resulted in 190 additional results. Note that in this paper, we adopted Rasch as an umbrella term that includes Rasch and the 1-parameter logistic (1PL) model.

After removing duplicate results, the search resulted in 169 pertinent articles. Given the search results, we employed the following inclusion and exclusion criteria to select relevant works for detailed review.

## Eligibility Criteria

The inclusion and exclusion criteria for studies are as follows:

1. *Study design*: There was no strict study design required in this review. Reviews regarding the IRT-derived ICF evaluation tool were considered, languages: “English”, conference abstracts without the full text, and unpublished studies were excluded;
2. *Participant characteristics*: Participants irrespective of gender, age, and health conditions were evaluated via the ICF categories;
3. *Outcome measures*: The outcome measure or self-report questionnaire include ICF items in body structure, body function, activity and participation, and environmental factors.

## Results

Our approach resulted in the identification of 46 publications that met all predetermined criteria (Figure 1). From each article, we extracted the information on several key variables, including the specific ICF core set, the original number of ICF categories, the number of IRT-based ICF categories, sample size, settings, response options, collapse strategy, scoring strategy, and data analysis software, which were summarized in Table 1, Table 2, Table 3 and Table 4.

**Figure 1.**
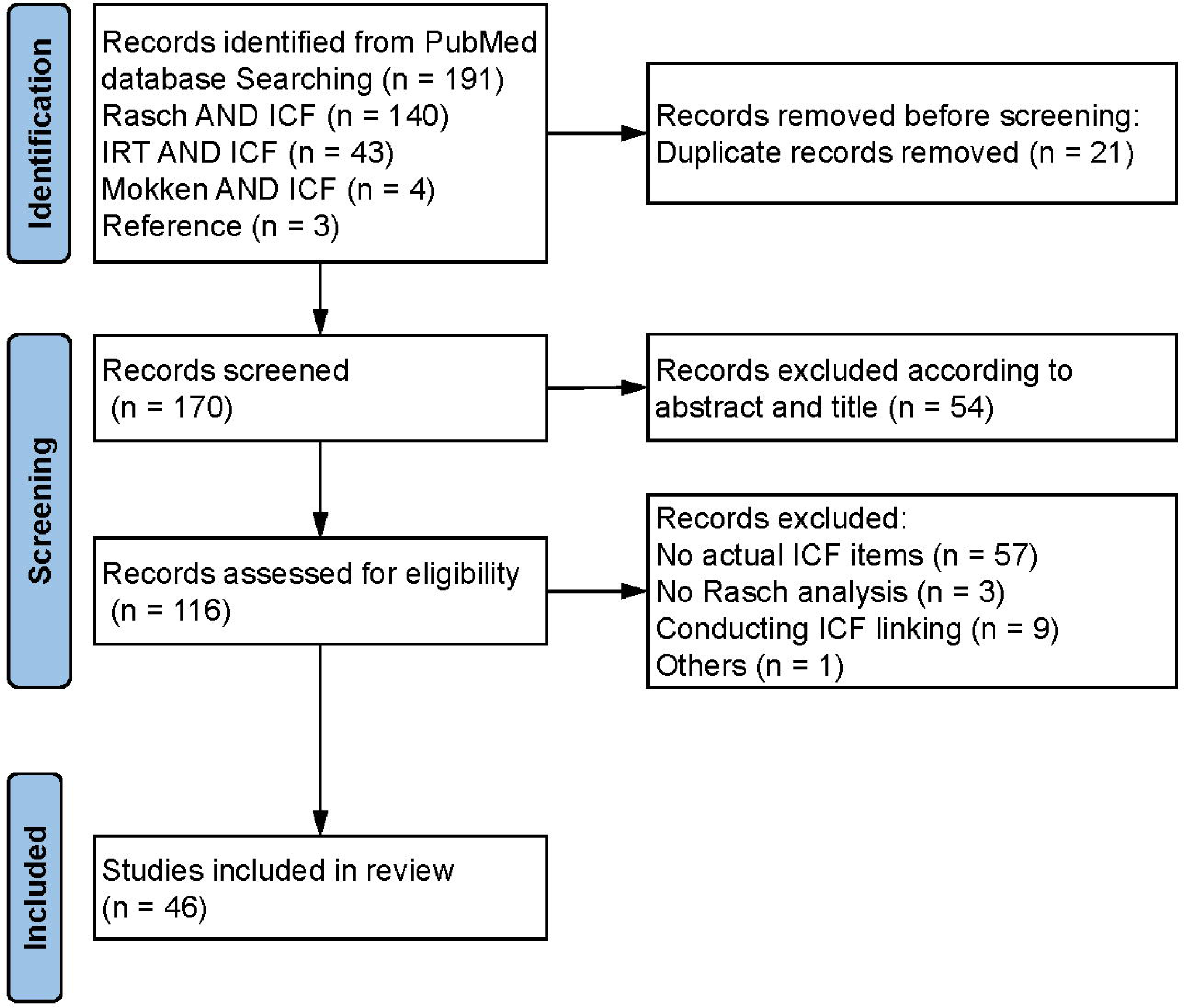
The Flow diagram of study selection.

**Table 1.**
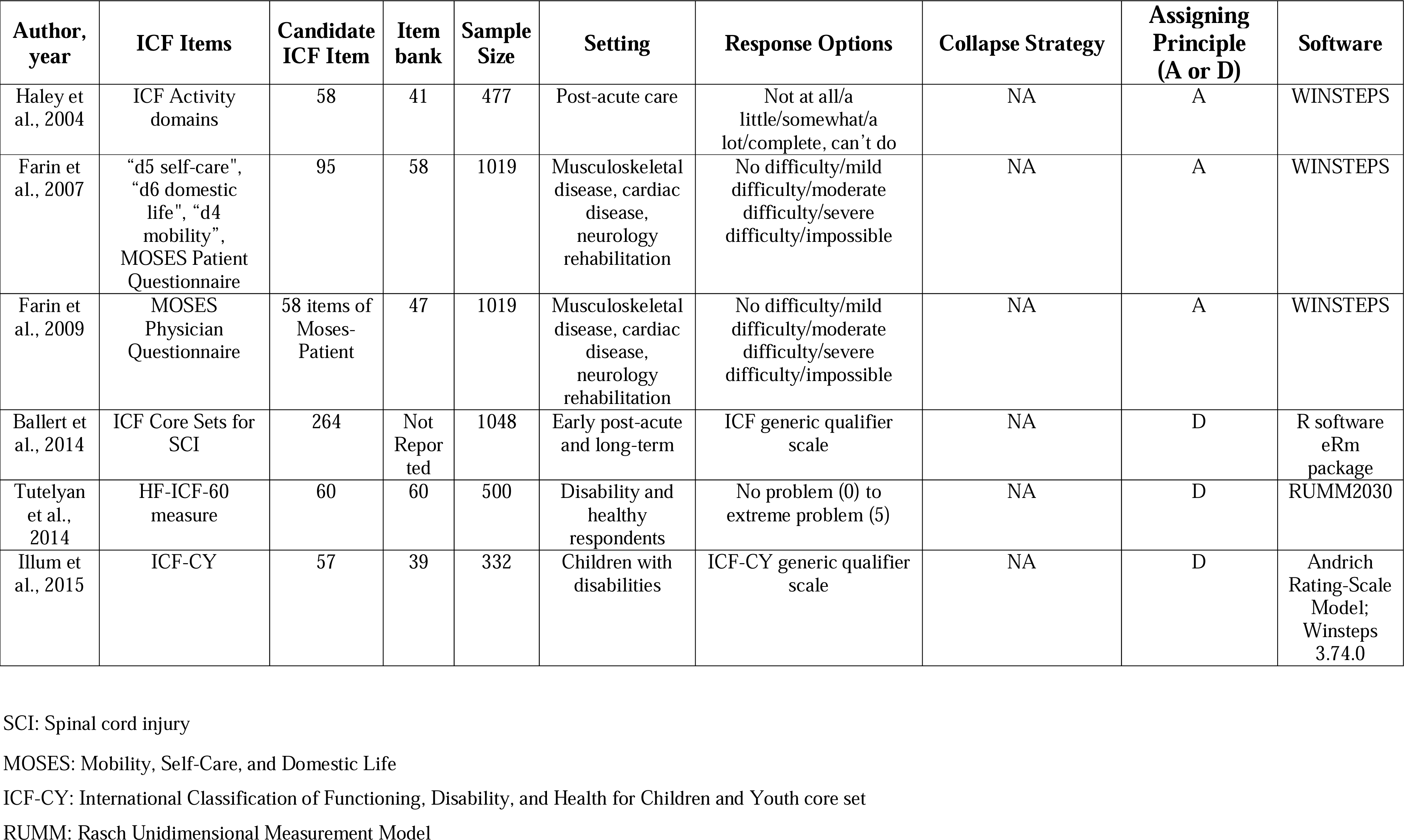
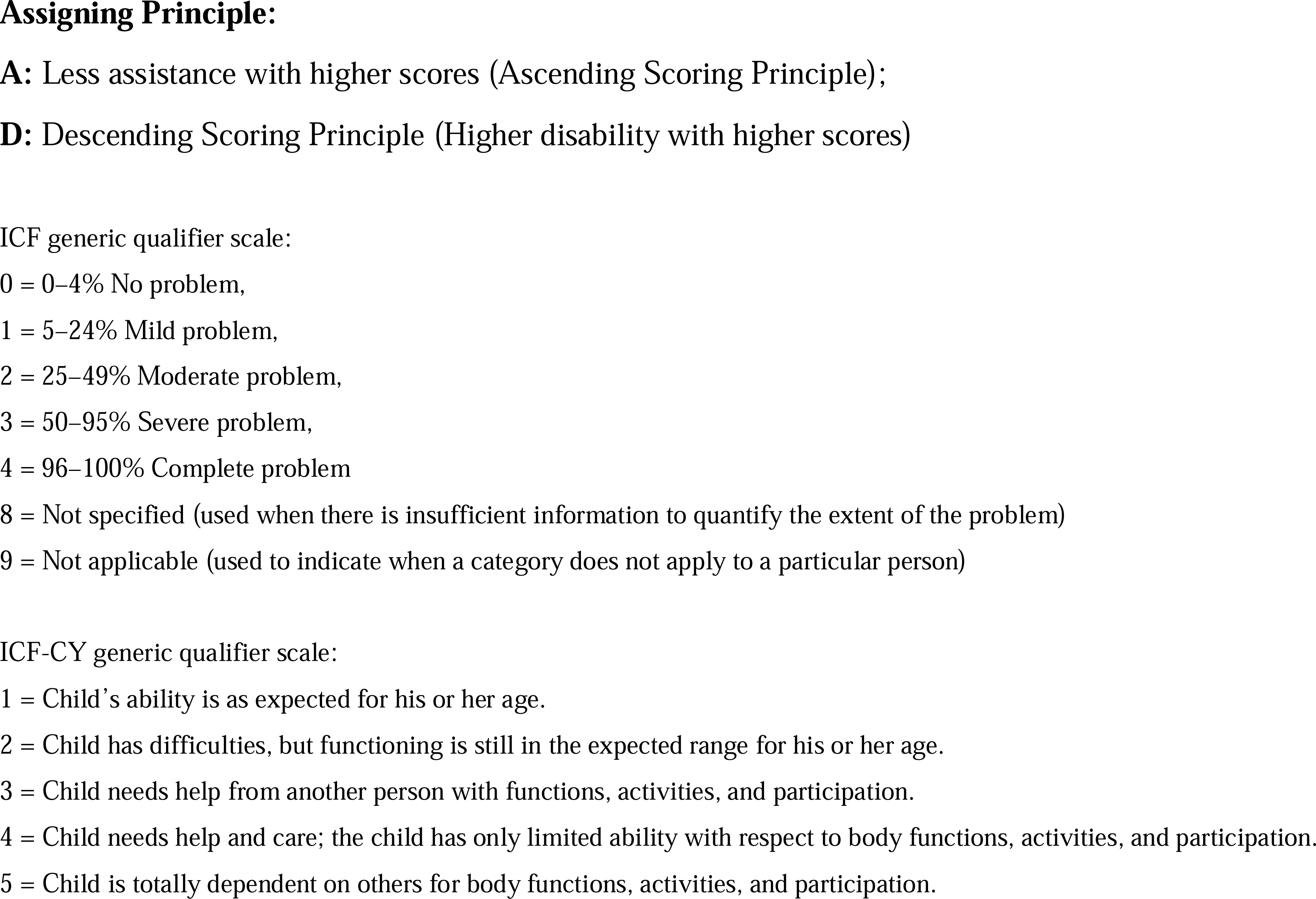
Dense Stratification Principle and more items based on the ICF-based IRT modeling.

## Sufficiency and Efficiency in Clinical Evaluation: Challenges of Operationalizing the ICF Questionnaire

The ICF provides a comprehensive taxonomy of function and disability, organized into four components, namely body structure, body functions, activity and participation, and environmental and personal contextual factors. However, the ICF functional category bank includes nearly 1,500 categories, making it impractical to assess each category in clinical practice. To address this challenge, the derivatives of ICF known as the ICF core set have been developed to evaluate functioning performance for specific health conditions or healthcare settings(Selb et al, 2015). There are two versions of the ICF core set: the comprehensive core set and the brief core set. A comprehensive ICF core set is inclined to exhaustively describe disease-specific functions, while it overloads the burden of answering lengthy and multi-item questionnaires on patients and clinicians. On the other hand, a brief ICF core set sacrifices the sufficiency of the questionnaire to satisfy the minimal standard of assessing the most essential functional categories. Despite the advantages of both core sets, two main issues are faced with the operationalization of the ICF questionnaire, namely the complex scoring system and data analysis in the ICF Likert scale.

Firstly, the ICF category in the core set requires scoring on a 0-4 Likert scale, which might not be an efficient application in real clinical scenarios. There are controversies regarding how to apply the 5-level Likert scale to measure the degree of “disability” regarding each ICF category, namely, “no impairment” = 0, “mild impairment” = 1, “moderate impairment” = 2, “severe impairment” = 3, and “complete impairment” = 4(Rauch, Cieza, and Stucki, 2008; Jette, 2018). Practitioners have not achieved full consensus on ICF category descriptions and differentiation between scale points, especially the qualifiers 2 to 4. Although researchers have suggested the employment of other assessment tools to assist in determining qualifier values, it significantly increases the complexity of the assessment process after expanding one questionnaire into multiple questionnaires. It will greatly expand the evaluation time due to a five-point rating scale for each item in either comprehensive or brief core sets. Thorough scoring options and extensive categories qualify the sufficiency of clinical assessment, but it is hard to satisfy the efficiency of evaluation in rapid-paced and hectic clinical work.

Another prominent barrier is the data analysis of the ICF Likert scale. Firstly, the summated scores represent the level of dysfunction depending on the uneven interval of the 5-point qualifier scale, which diminishes the inter- and intra-individual comparability of outcome measures. Secondly, each ICF category is considered as equal difficulty in the daily assessment rather than hierarchically categorize in terms of parameters such as item difficulty and discrimination. Without considering the quantitative values for item difficulties regarding certain individual abilities, simple qualifier values of each item are insufficient to represent the whole picture of clinical outcomes.

## Personal Ability and Item Difficulty: Principles and Values of the IRT-derived Medical Outcome Measures

Chang et al.(1995) proposed that the IRT could be a promising tool for measuring health outcomes and quantifying the degree of health, which has been applied in the medical field since the early 2000s(Hays, Morales, and Reise, 2000). The IRT model differentiates between an individual’s ability levels and the difficulty levels of test items, constructing an Item Response Function (IRF) by taking θ as an independent variable and the probability of correct answer rate (P(θ)) as a dependent variable, under the condition that test items do not interact with each other. For example, Liu et al.(2002) conducted a Rasch analysis on the Stroke Impairment Assessment Set (SIAS). After reducing subsets and combining similar items, the IRT-derived SIAS is not only transformed into a linear scale but also can reflect person abilities corresponding to item difficulties.

In recent years, there has been increasing interest in the use of IRT-based Rasch models for processing ICF evaluation data, which can be broadly categorized into two perspectives. The first approach involves handling ICF categories as questionnaire items, which has been used in various studies (Table 1, Table 2, Table 3, Table 4), such as the development of a 41-item activity measurement scale for post-acute care by Haley et al.(2004). Cieza et al.(2009b) also constructed the Rasch modeling based on the comprehensive assessment of the functional level among patients with rheumatoid arthritis (AS) and osteoarthritis (OA). By investigating 437 patients with OA in Germany, Italy, Hungary, Serbia, and Singapore, researchers achieved the first ICF-based cross-cultural clinical functional measurement. They found that the difficulty of the same ICF category varied in different countries. The Rasch analysis also explored ICF categories for various conditions or diseases, including AS(Cieza et al, 2009a), OA(Cieza et al, 2009b; Kurtaiş et al, 2011), chronic widespread pain(Prodinger et al, 2012), low back pain(Bagraith, Strong, Meredith, and McPhail, 2017; Røe et al, 2009; Røe, Bautz-Holter, and Cieza, 2013), breast cancer(Yang, Shin, Shin, and Lim, 2014), children and youth with disabilities(Illum and Gradel, 2015), the ICF Generic Set(Ehrmann et al, 2018), the Rehabilitation Set(Gao et al, 2020), spinal cord injury (SCI)(Li et al, 2018; Jia et al, 2020), stroke(Feng, Jiang, Sun, and Lin, 2022), haemophilia(Feng et al, 2022), and cerebral palsy(Jiang et al, 2023).

The second approach involves linking questionnaire items from scales to ICF items, which has been applied in several studies(Finch, Higgins, Wood-Dauphinee, and Mayo, 2009; Maritz et al, 2022; Nilsson, Westergren, Carlsson, and Hagell, 2010, 2010; Prodinger, O’Connor, Stucki, and Tennant, 2017; Van de Winckel et al, 2019). Finch et al.(2009) discerned 44 items from other scales and constructed a Rasch model based on the ICF framework to evaluate the level of physical function in patients three months post-stroke. They obtained an item difficulty list consisting of 52 items, and suggested that stroke rehabilitation goals and treatment plans could be developed based on person abilities and item difficulties. This approach has been shown to be useful for identifying functional limitations and developing interventions that target specific areas of impairment. Other studies have used similar approaches to link scales to the ICF framework for various health conditions, including Parkinson’s Disease(Nilsson, Westergren, Carlsson, and Hagell, 2010), stroke(Van de Winckel et al, 2019), spinal cord injury(Maritz et al, 2022), demonstrating the versatility and applicability of this method in the assessment of functional outcomes.

## Research Progress and Deficiency: Clarification of the ICF-based IRT Scoring Principles

ICF-based IRT studies employ two dimensions of scoring principles, stratification and assignment. The stratification dimension involves sparse and dense stratification principles to grade each ICF category. The sparse stratification principle (SSP) can involve scoring each ICF category with qualifiers 0 for ‘no impairment/limitation’, 1 for ‘impairment/limitation’ instead of adopting the original 5-level Likert scale(2005), or utilizing collapse strategies, item combination, and item deletion during data analysis. Some studies suggest that reducing the five-point to three-point qualifiers leads to a better model fit. For example, Li et al. (2018) found that reducing the five-point (01234) to the three-point (01112) qualifiers improves the overall model fit. Gao et al.(2020) also recommend using a collapse strategy and converting from 01234 to 01223 or 01112 in the Rasch model when operating the ICF five-point qualifiers as a rating scale. On the other hand, the dense stratification principle (DSP) expands qualifiers to capture more detailed clinical information and reduce the ceiling and floor effect, using an 11-point numeric rating scale and transforming raw scores to the Rasch-based interval score(Ehrmann et al, 2018). The SSP is broadly endorsed for the qualifiers in terms of enough items, such as 65 ICF categories of SCI(Li et al, 2018). The DSP is more comprehensive but requires fewer items, such as the Generic 6 set(Ehrmann et al, 2018) in Table 2.

**Table 2.** Dense Stratification Principle and Less Items based on the ICF-based IRT Modeling.

| Author, year | ICF Items | Candidate ICF Item | Item bank | Sample Size | Setting | Response Options | Collapse Strategy | Assigning Principle (A or D) | Software |
| --- | --- | --- | --- | --- | --- | --- | --- | --- | --- |
| Prodinger et al., 2012 | Brief ICF Core Set for fibromyalgia | 19 | 14 | 256 | Patients with Fibromyalgia | ICF generic qualifier scale | 01123 | D | RUMM 2020 |
| Reinhardt et al., 2016 | ICF Generic Set categories | 7 | 7 | 761 | Musculoskeletal, neurological health condition, SCI and TBI |  | NA | D | RUMM 2030 |
| Illum and Gradel, 2017 | ICF-CY | 26 | 26 | 162 | Parents of children with disability |  | NA | D | Winsteps 3.74.0 |
| Illum et al., 2018 | ICF-CY | 26 | 26 | 162 | Parents of children with disability |  | NA | D | Winsteps 3.74.0 |
| Reinhardt et al., 2016 | ICF Generic Set categories | 7 | 7 | 761 | Musculoskeletal, Neurological health condition |  | NA | D | RUMM 2030 |
| Guo et al., 2020 | Activity and participation categories in the brief ICF core set for COPD | 4 | 4 | 103 | Patients with COPD |  | NA | D | FACETS (Minifac) 3.67.0 |
| Alviar et al., 2012 | ICF core set for OA | 32 | 25 | 316 | Adults with OA who were either waiting for hip or knee arthroplasty (pre-operative), or within one year of surgery (post-operative) |  | 01123 | D | RUMM 2030 |
| Jia et al., 2020 | SCI-related ICF items | 31 | 28 | 112 | Families of SCI patients |  | NA | D | RUMM 2030 |
| Misajon et al., 2008 | Self-care, Mobility, Participation, Relationships, and Psychological Well-being | 23 | 23 | 210 | Mobility impairment | Impact scale: “no impact” to “extreme impact”, or Distress scale: “no distress” to “extreme distress”; 6-point scale (012345) | 011223 | D | RUMM 2020 |
| Pallant et al., 2009 |  | 23 | 22 | 259 | Patients with OA |  | NA | D | RUMM 2020 |
| Prodinger et al., 2019 | Mobility, Selfcare, Domestic life | 25 | Not reported | 2927 | Stroke, RA, OA | No problem to complete problem; 5-point scale (0-4) | NA | D | RUMM 2030 |
| Janeslätt et al., 2015 | Time-Self rating ICF environmental factors | 5 | 5 | 64 | Adults with/without mental disability | Not at all/a little/a lot/very much (1234) | NA | A | Winsteps |
| Prodinger et al., 2016 | SCIM-SR | 17 | 17 | 1549 | Traumatic or nontraumatic SCI | Total assistance/partial assistance/independent but need adaptive devices or specific equipment/ independent and do not need adaptive devices or specific equipment | NA | A | RUMM 2030 |

SCIM-SR: Spinal Cord Independence Measure – Self Report; TBI: traumatic brain injury ; COPD: chronic obstructive pulmonary disease; RA: rheumatoid arthritis; OA: Osteoarthritis

Considering assignment dimension, most ICF-based IRT studies have typically employed the “descending assignment principle or assigning by dysfunction (ABD)” to address item scores based on functional ability, where lower functionality corresponds to a higher score. Some examples of studies that have employed this principle include Farin et al.(2007), Cieza et al.(2009a), Røe et al.(2013), Ehrmann et al.(2018), Guo et al. (2020), and Gao et al.(2020). However, a few studies have taken an alternative approach, using the “ascending assignment principle or assigning by functioning (ABF)”, where a higher score corresponds to higher functionality(Feng et al, 2022; Feng, Jiang, Sun, and Lin, 2022; Jiang et al, 2023). Okochi et al.(2013) implemented the binary response options, scoring items as “yes” for functioning (assignment = 1), or “no” for dysfunction (assignment = 0). According to the final scores, they classified individuals into functional stages based on their activity level. The Lucerne ICF-based Multidisciplinary Observation Scale (LIMOS) also adopted the ascending assignment principle by generating 45 ICF-similar items based on the degree of assistance in daily life(Van de Winckel et al, 2019).

## Discussion

To achieve an optimal balance between sufficiency and efficiency in clinical assessment, it is imperative to develop a concise and effective core set from the larger and more comprehensive ICF core set. This core set should contain minimal yet discriminative items, capturing continuous functioning changes, and ensuring good construct validity and internal consistency. The implementation of the IRT statistical approach has been used to derive such a measurement tool, which we refer to as the ICF parsimonious core set (PCS).

By overcoming the limitation of assuming that each functional item is equally difficult, the IRT modeling approach has proven to be an effective way to extract PCSs. It has been applied since the inception of the ICF and has demonstrated its effectiveness over the last two decades. The IRT-derived outcome measure has the potential to serve as a pragmatic tool that provides comprehensive and holistic information that can facilitate decision-making in clinical settings.

## Future Implication: the ICF Framework and IRT Technique

As the most commonly used IRT model, the Rasch analysis can optimize clinical scales due to its ability to enhance the construct validity of clinical scales and reduce the indiscriminative items. Based on different configurations of parameters, the IRF can be presented as an S-shaped Item Characteristic Curve (ICC, Figure 2) on the coordinate axis. This statistical model overcomes the limitation of the classical test model, which assumes that each item is equally difficult. The IRT recognizes that item difficulty is represented by parameters and maps the locations of individuals and the difficulties of test items onto the same invariant scale, allowing direct comparison. The IRT-based measurement of individual ability has important implications for clinical assessment in rehabilitation practice. It can help identify an individual’s functional level compared to others with the same disease, as well as assess whether a specific function has improved following intervention. Additionally, the IRT-based estimation of item difficulty can aid in prioritizing rehabilitation goals by determining which functional items are most appropriate for an individual’s abilities and which dysfunctions should be addressed first.

**Figure 2.**
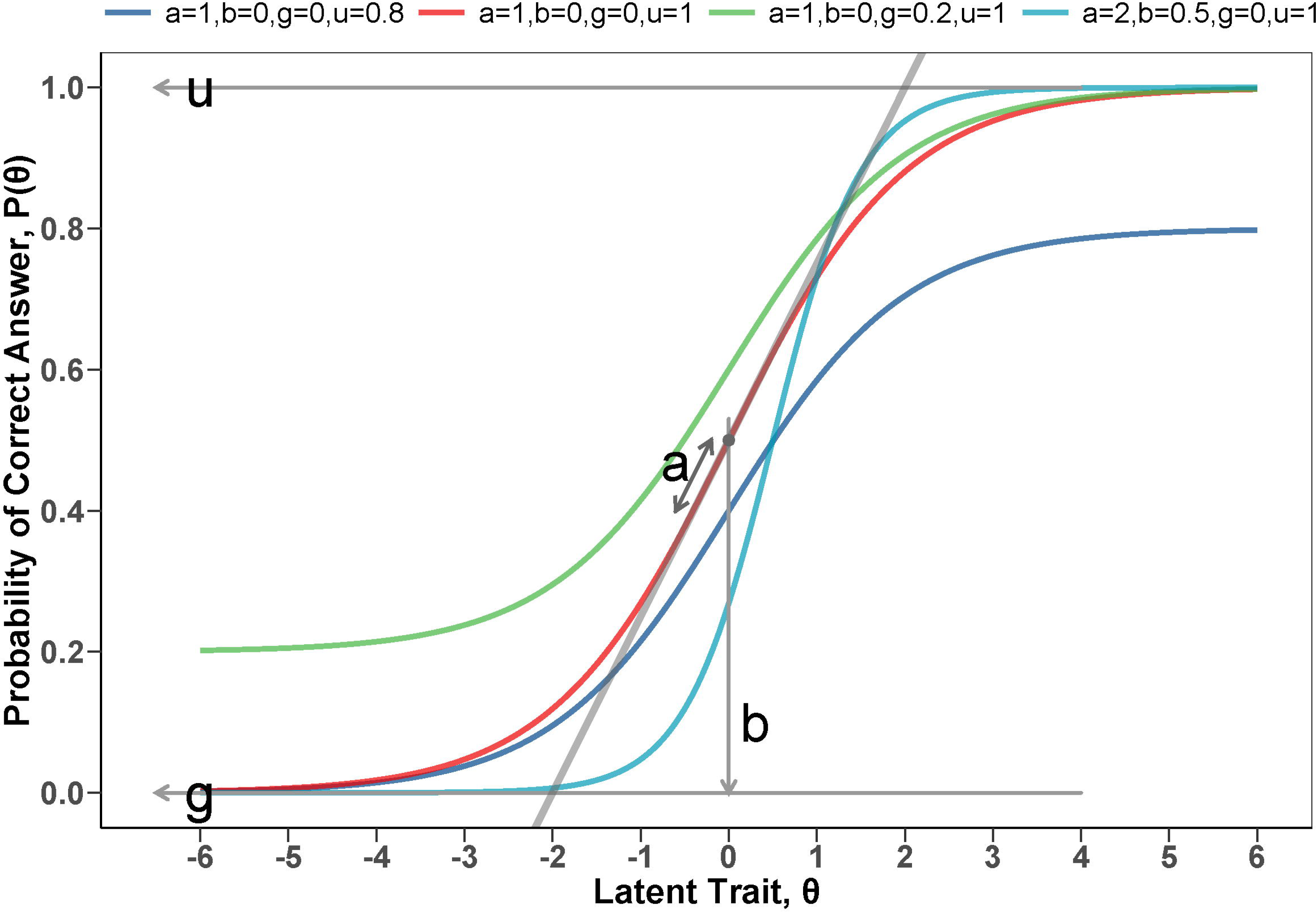
Schematic diagram of the ICC curve. The illustration of the effect of four parameter were respectively depicted on the shape of the ICC (a = discrimination/slope, b = difficulty/location/threshold, g = pseudo-guessing/lower asymptote, u = upper asymptote/slipping/pseudo-carelessness)

Supplemental Table 1 displays four parameters within the IRT model, where the parameter “difficulty (d)” is primarily reserved in parametric IRT models. As an illustration, the Rasch model only estimates the “difficulty” parameter, and uses default values of a = 1 (moderate discrimination), g = 0 (no guess), and u = 1 (completely carefulness). The parameters θ and d can be expressed numerically on the same scale, namely logit or the log-odds (the logarithm of the frequency ratio of scored and non-scored items). Consequently, individual functional status can be sorted by person abilities, and different items can be hierarchized according to item difficulties. Moreover, person abilities and item difficulties can be compared on the same scale, allowing for the identification of whether an item is relatively difficult (θ < d) or relatively easy (θ > d) for a person with a certain ability level (θ). The IRT model can also convert an ICF Likert scale with ordinal response options into an equidistant scale(Prodinger et al, 2020).

The parameters, g and u, offer an opportunity to identify and address issues with the quality of the ICF assessment scale. For instance, if a model reports an item with high guessing or carelessness, the evaluators or clinical practices in the institution should audit certain functioning evaluation processes to improve the assessment quality.

## The Scoring Principle: Sparse Scoring principle and Assigning by Functioning

The SSP and DSP principles are useful in different contexts, with the SSP being more efficient and the DSP being more comprehensive, as shown in Figure 3. From an efficiency perspective, the stratification principle of qualifiers aligns with the SSP, such as screening tools (Table 3). In contrast, research tools with a large number of items and DSP can capture functional limitations comprehensively. To strike a balance between sufficiency and efficiency, it is recommended to adopt more items in ICF core sets, while using SSP for detailed assessments (Table 4). A recent study by Feng et al.(2022) extracted 118 categories of “b” and “d” components from the ICF categories for stroke and used a dichotomous qualifying strategy (dysfunction = 0 or functioning = 1) for IRT modeling. The final Rasch-derived ICF scale consisted of 45 items. This approach enabled simplified assessment of comprehensive ability levels and allowed for scoring of functional categories based on ICF qualifier scores. By utilizing a binary response pattern, the approach may further enhance efficiency.

**Figure 3.**
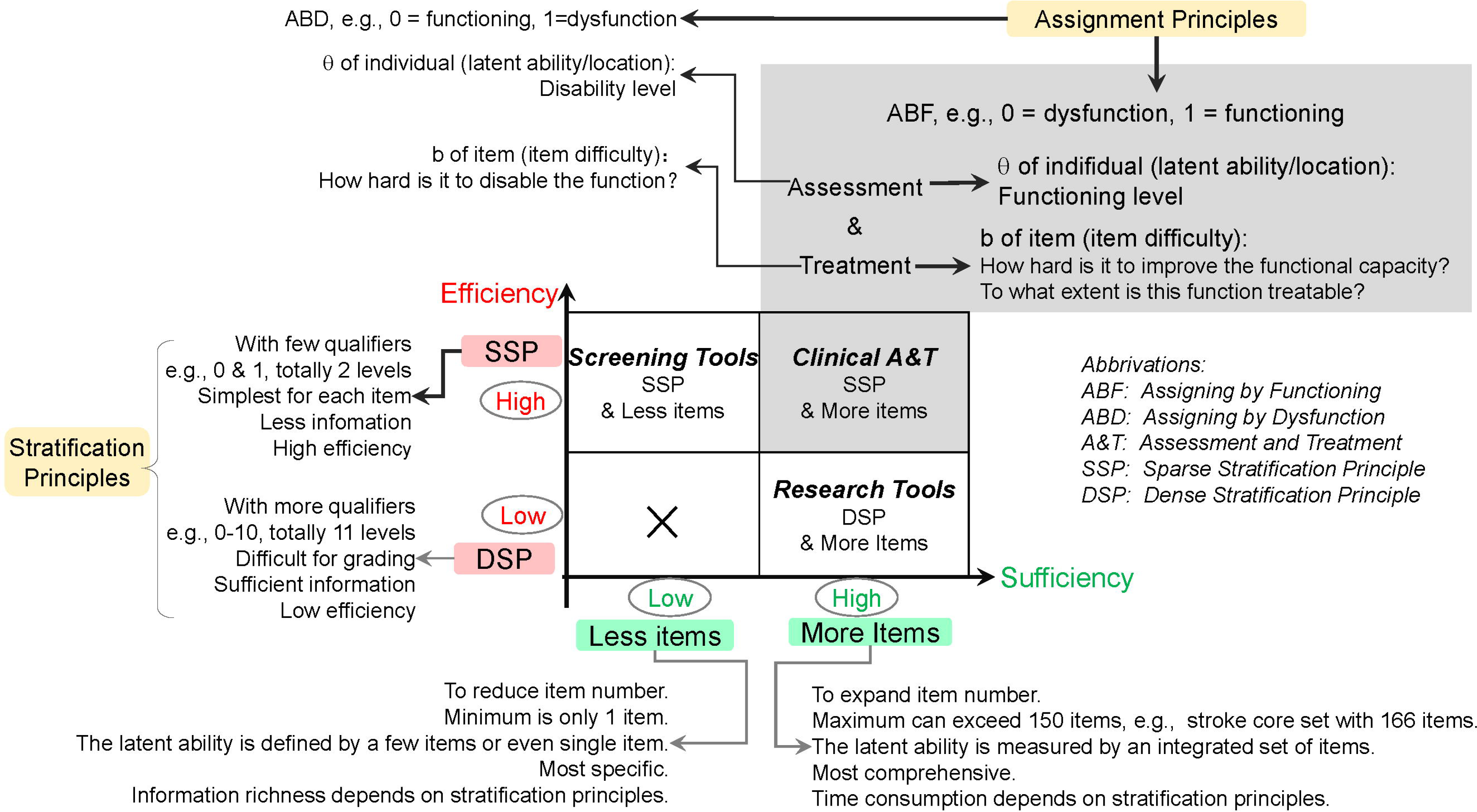
Exploration of efficiency and sufficiency in the research paradigm of the ICF-based evaluation tools.

**Table 3.**
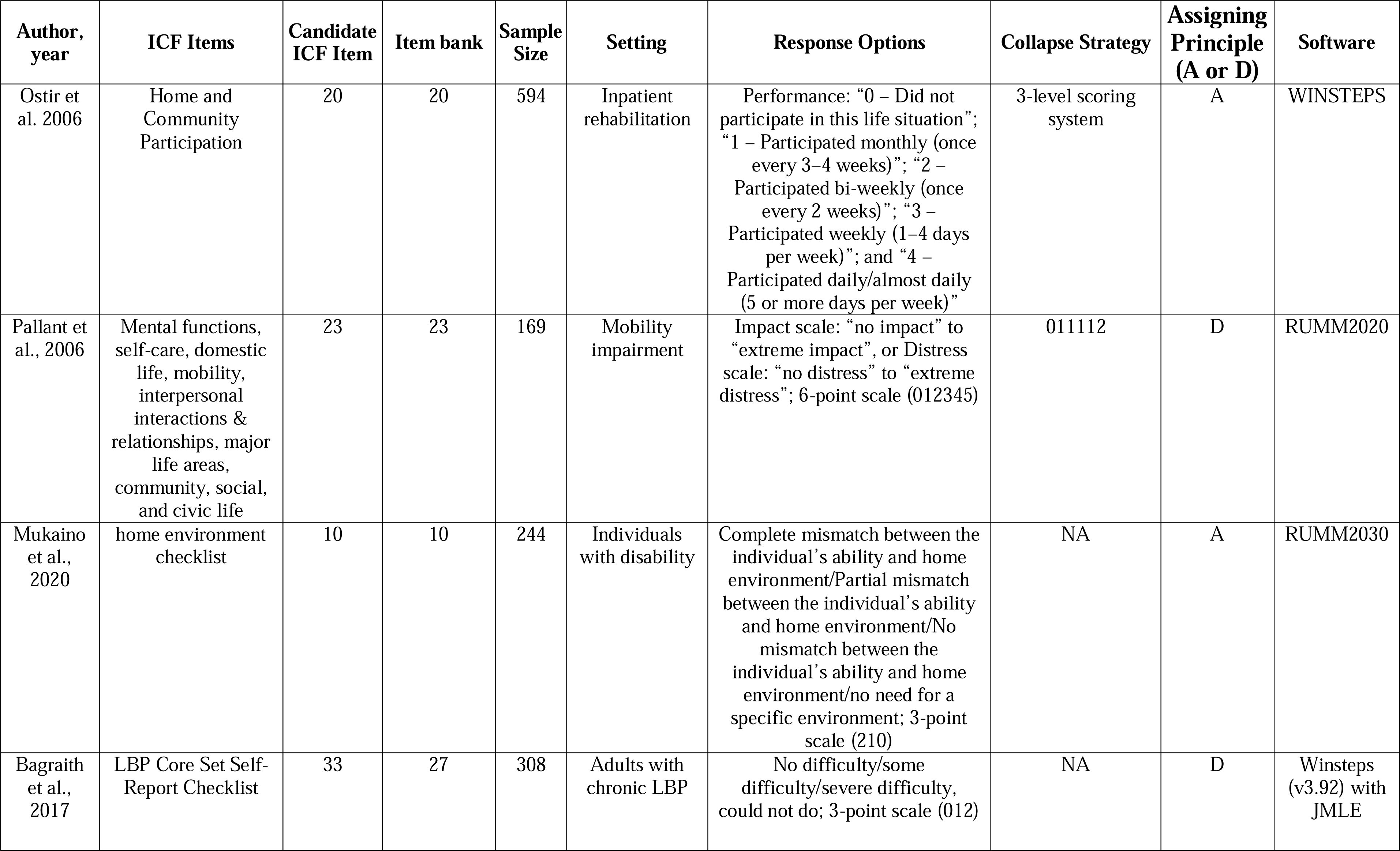
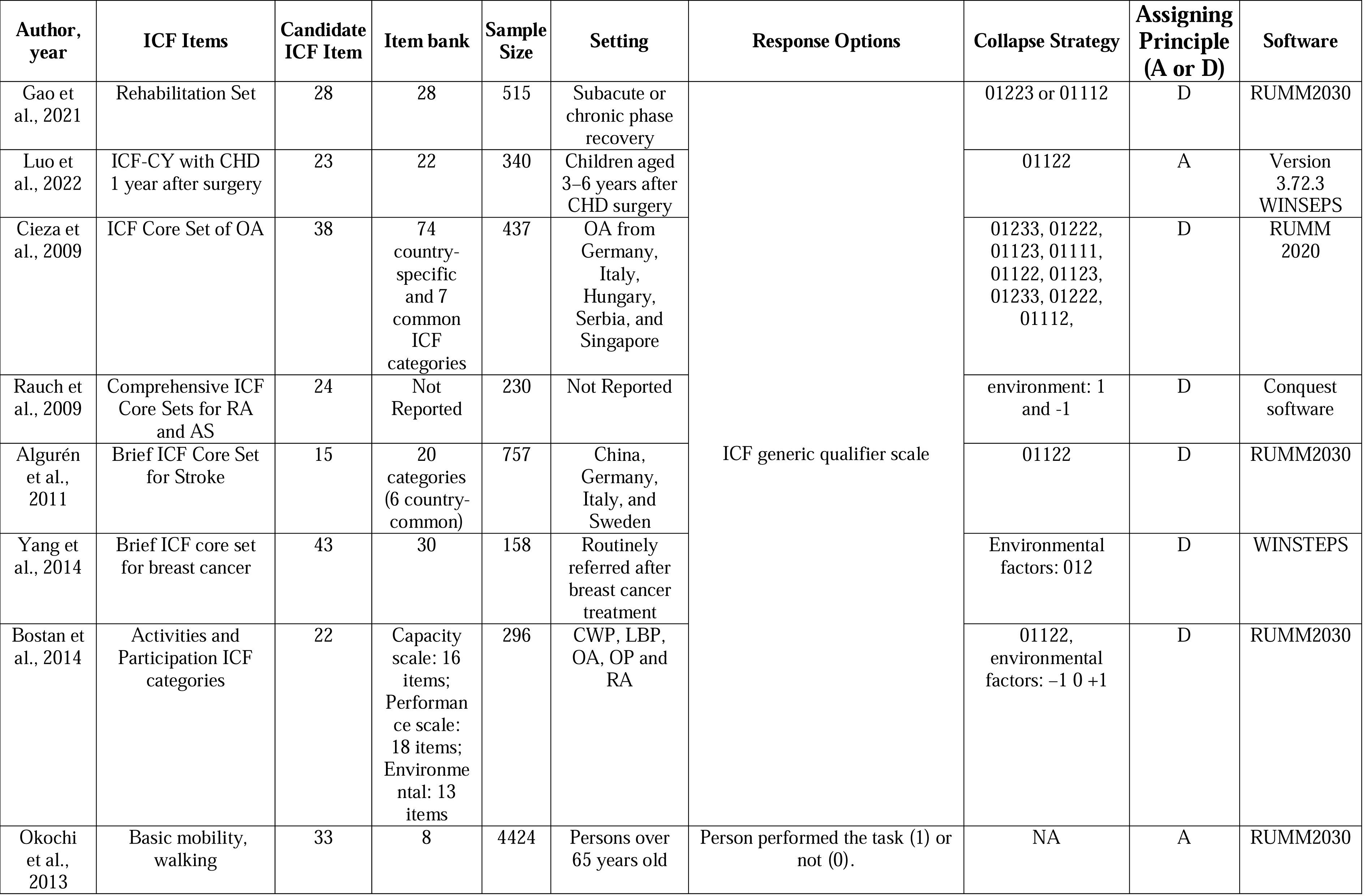
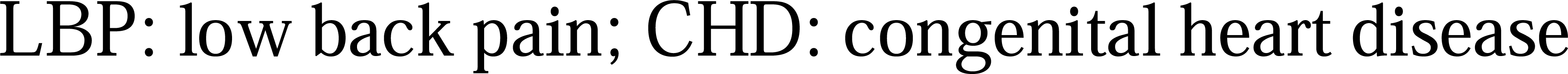
Sparse Stratification Principle and Less Items based on the ICF-based IRT Modeling.

**Table 4.**
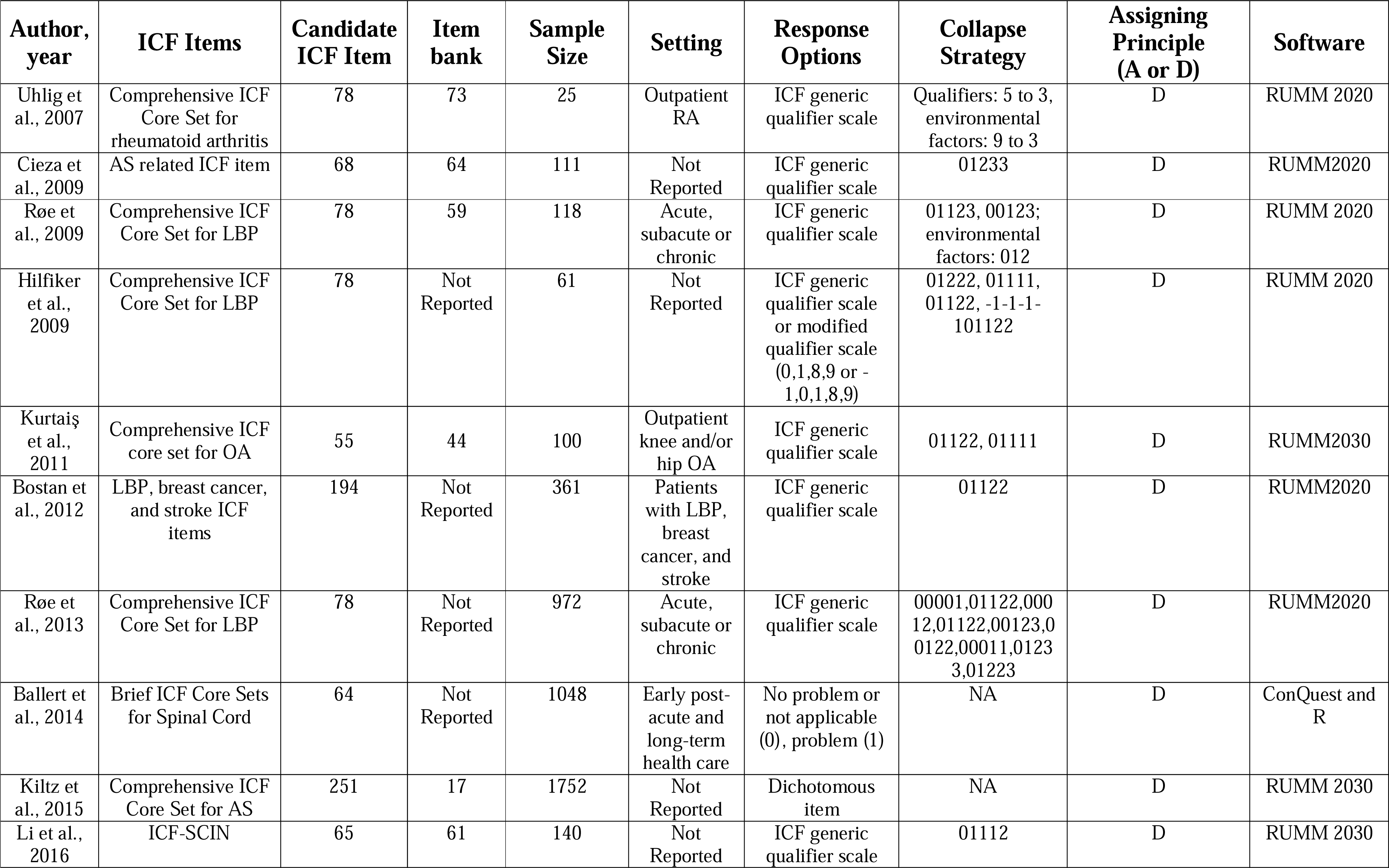
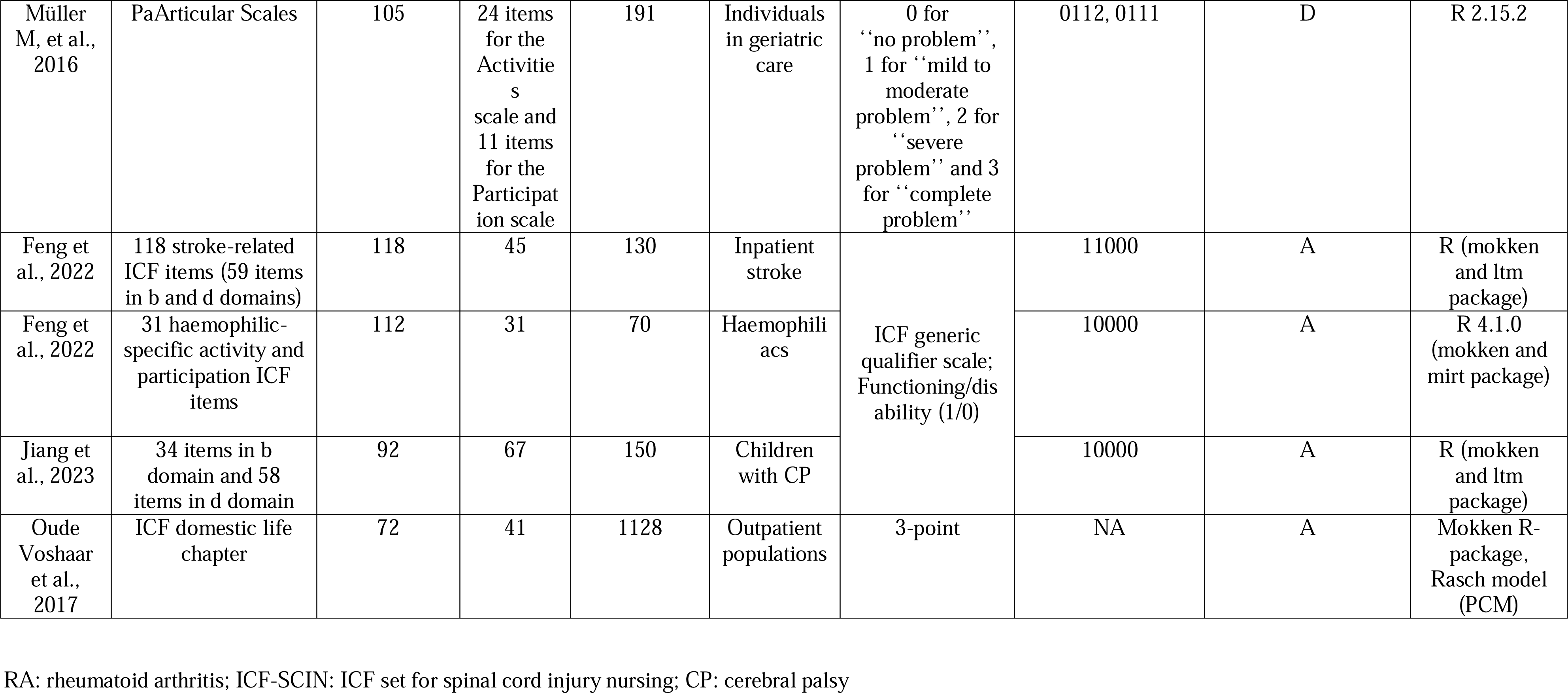
Sparse Stratification Principle and More Items based on the ICF-based IRT Modeling.

In terms of assignment scoring principles, the ABD principle and the ABF principle have no difference in assessing an individual’s functional capacity. A reduction in dysfunction or an improvement in functional level is typically considered an improvement result. However, differences between the two principles become evident in IRT analyses and guiding treatment plans. The logit value of item difficulty is the primary contradiction of evaluation and treatment in rehabilitation services. The ABD principle presents a semantic dilemma in clinical thinking as it indicates how difficult it is to disable a certain function. On the other hand, the ABF principle aligns with the clinical treatment thinking process of improving functional capacity, and only the logit value of an individual’s potential traits derived from the principle of ABF would reflect the level of individual function. As assessment is in the service of treatment, researchers have recommended adopting the ABF principle in future ICF studies(Feng, Jiang, Sun, and Lin, 2022; Feng et al, 2022).

## The IRT-derived ICF Parsimonious Core Set and Wright Map

The ICF-based IRT questionnaire, constructed with the principles of sufficiency and efficiency, uses a binary response pattern to increase efficiency. ABF scoring aligns with difficulty parameter in the IRT modeling, and the resulting IRT-derived PCS has high internal consistency and construct validity. This ICF PCS contains the most discriminable and minimal items, making it ideal for a clinical-pragmatic approach. Empirical evidence from PCS measurement tools, such as the stroke simplified IRT assessment tool(Feng, Jiang, Sun, and Lin, 2022), IRT-derived and ICF-based haemophiliac activity and participation assessment tool(Feng et al, 2022), and ICF-based IRT assessment tool(Jiang et al, 2023) for children with cerebral palsy, demonstrates its efficacy in assessing functional stages.

A novel approach to prioritizing daily activities intervention can be achieved by understanding the difficulty levels of diverse functional activities. The Wright map, which plots person abilities and item difficulties with the same unit (i.e., logit), orders the level of functional ability of participants on the left side and the difficulty of functional categories on the right side (Figure 4 © 2022 John Wiley & Sons Ltd.). The most difficult functional categories are at the top of the scale, while the easiest ones are at the bottom. Subjects with low functional ability struggle with even the easiest activities, while those with higher functional ability experience no difficulty performing body functions or activities.

**Figure 4.**
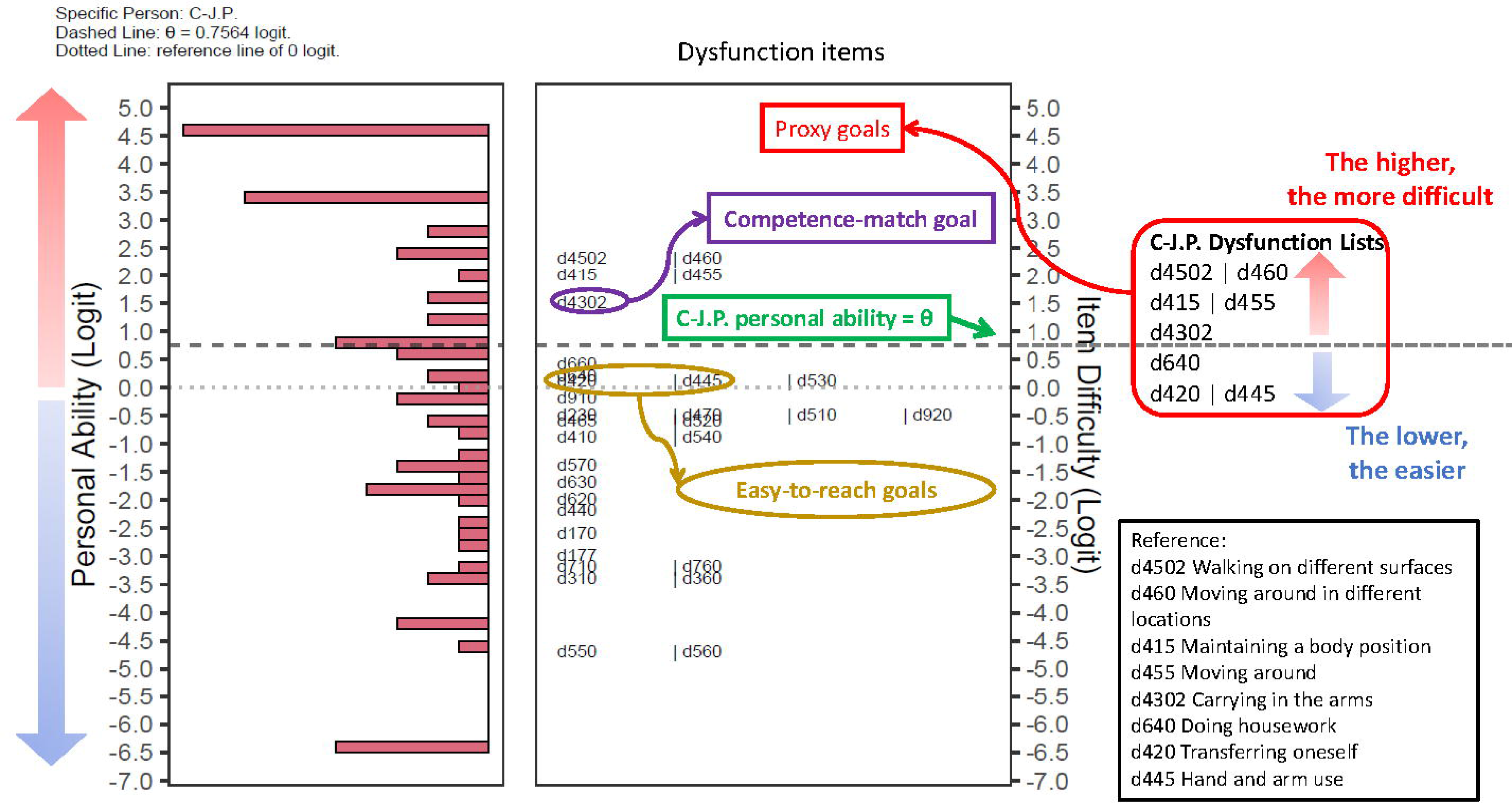
Personal ability and item difficulty constructed in the Wright map. The left column illustrates the personal ability corresponding to the item difficulty in the right column © 2022 John Wiley & Sons Ltd.

Recent advancements in technology and data analytics enable the use of IRT-derived PCS functioning outcome measures to personalize interventions that improve functioning and quality of life. The Wright map provides evidence for customized rehabilitation goals by integrating person abilities and item difficulties. Item difficulties can be used to estimate rehabilitation goals, such as the level of difficulty in improving a specific function from a low stage to a high stage. Additionally, several functioning properties can provide more proxy goals for rehabilitation therapy.

## Conclusion

The use of the IRT technique in ICF studies has highlighted a number of contradictions, such as the trade-offs between efficiency and adequacy, comprehensiveness and precision, assessment and treatment, and disability and health. In the context of humanism in healthcare, the fundamental goal of function evaluation and treatment intervention must be to optimize patient outcomes. To achieve this, we can consider the following suggestions through comparative analysis: (1) Streamline the stratification of qualifiers to achieve efficient evaluation, while increasing the number of scale items to ensure comprehensive assessment; (2) Adhere to the principle of “assigning by functioning” when scoring items to promote clinical-relevant and intuitive thinking in tailoring treatment plans; (3) Adoption of the Wright map (person-item threshold map) might facilitate patient-oriented care according to person abilities and item difficulties.

## Acknowledgment

## Data Availability

Data sharing is not applicable to this article as no datasets were generated or analyzed during the current study.

## Acknowledgements

Not applicable.

## Ethical Approval and Consent to Participate

Not applicable.

## Data Availability Statement

Data sharing not applicable to this article as no datasets were generated or analyzed during the current study.

## Author contribution

CF and LF conceived the original idea. CF and LF designed and performed the review and wrote the manuscript. LF offered essential idea to the review design. CF and LF edited the manuscript. SGL gave comments on the review design. All authors discussed the conclusions and commented on the manuscript.

