## Supplemental Table 1 for "Research Paradigm of the International Classification of Functioning, Disability and Health (ICF) with Item Response Theory: Clarification, Classification, and Challenge"

Supplemental Table 1 Four parameters of parametric IRT model

| Parameter | Terminology | Explanation |
| --- | --- | --- |
| a | discrimination/ slope | As the slope line with two-side arrows along with the red curve depicted in Figure 2, the item discrimination is the slope value at the inflection point of the item characteristic curve (ICC). The test scores will be most affected by the variation of person abilities at this inflection point. Accordingly, a slight change in the individual ability will greatly impact the item scores, so as to hierarchize examinees’ ability levels. |
| b | difficulty/ location/ threshold | The vertical arrow across the inflection point in Figure 2 pointing vertically to the horizontal axis demonstrates the latent trait (θ) of the ICC, that is, the “location” of the horizontal axis at the point of maximum slope. The ICC maintains its shape and shifts to the left or right with the changes in the b value. As the b value increases, the ICC curve will shift to the right. It will lead to a decrease in the scoring rate of the test items, even though person abilities remain unchanged, indicating that the item difficulties increase. Instead, when the b value decreases, the ICC curve moves to the left, and item difficulties decrease. For the 2-parameter logistic (2PL) model, the b value can be converted to easiness (the intercept parameter denoted by “d”) through the following equation: *d=-ab*. |
| g | pseudo-guessing/  lower asymptote | The lower arrow pointing horizontally to the left vertical axis shown in Figure 2 is the lower limit of the ICC, that is, the lowest accuracy rate that items can achieve. It emphasizes the possibility that guesses will result in getting a point (false score). The ideal test performance would be zero guesses, meaning that examinees with low person abilities would not be able to get a score by guessing. High guess possibility may underestimate item difficulties. |
| u | upper asymptote/ slipping/  pseudo-carelessness | The upper arrow pointing horizontally to the left vertical axis displayed in Figure 2 is the upper limit of the ICC, that is, the highest accuracy rate that items can achieve. It focuses on the possibility of losing points for an item due to carelessness. The ideal test performance would be zero carelessness or complete carefulness, meaning that examinees with high person abilities would likely score 100%. The low value of the upper asymptote may overestimate item difficulties. Quick assessments might lead to the lower item upper asymptote. |
